# Longitudinal microstructural MRI markers of demyelination and neurodegeneration in early relapsing-remitting multiple sclerosis: magnetisation transfer, water diffusion and g-ratio

**DOI:** 10.1101/2022.06.08.22276139

**Authors:** Elizabeth N. York, Rozanna Meijboom, Michael J. Thrippleton, Mark E. Bastin, Agniete Kampaite, Nicole White, Siddharthan Chandran, Adam D. Waldman, the FutureMS Consortium

**Author notes:** **Correspondence:** Elizabeth N. York, Adam D Waldman, Centre for Clinical Brain Sciences, University of Edinburgh, Edinburgh BioQuarter: Chancellors Building, Edinburgh EH16 4SB, United Kingdom.

## Abstract

**Introduction:** Quantitative microstructural MRI, such as myelin-sensitive magnetisation transfer ratio (MTR) or saturation (MTsat), axon-sensitive water diffusion Neurite Orientation Dispersion and Density Imaging (NODDI), and the derived aggregate g-ratio, may provide more specific markers of white matter integrity than conventional MRI for early patient stratification in relapsing-remitting multiple sclerosis (RRMS). The aim of this study is to determine the sensitivity of such markers to longitudinal pathological change within cerebral white matter lesions (WML) and normal-appearing white matter (NAWM) in recently diagnosed RRMS.

**Methods:** Seventy-nine people with recently diagnosed RRMS, from the FutureMS longitudinal cohort, were recruited to an extended MRI protocol at baseline and one year later. Twelve healthy volunteers received the same MRI protocol, repeated within two weeks. Ethics approval and written informed consent were obtained.

3T MRI included magnetisation transfer, and multi-shell diffusion-weighted imaging. NAWM and whole brain volume (WBV) were segmented from 3D T1-weighted MPRAGE, and WML from T2-weighted FLAIR. MTR, MTsat, NODDI isotropic (ISOVF) and intracellular (ICVF) volume fractions, and aggregate g-ratio were measured within WML and NAWM. G-ratio was calculated from MTsat and NODDI data.

Longitudinal change in brain volume and microstructural metrics was assessed with paired t-tests (α=0.05) and linear mixed models, corrected for confounding factors and multiple comparisons. Longitudinal changes were compared with test-retest Bland-Altman limits of agreement from healthy volunteer control white matter. The influence of varying MTsat and NODDI parameters on derived g-ratio was explored *in silico*.

**Results:** In NAWM, paired t-tests show a significant increase in g-ratio (p=0.012) and ICVF (p=0.025), and a decrease in MTsat (p=0.033) over one year, but no change in MTR, or ISOVF. Linear mixed models show effects in NAWM remain significant after adjusting for covariates and after False Discovery Rate (FDR) correction for multiple comparisons, and no significant change in WBV. In WML, MTsat and NODDI metrics increase significantly over one year (linear mixed models, FDR-corrected p<0.05). *In silico* simulations show that increase in g-ratio may result from a decrease in MTsat or ISOVF, or an increase in ICVF.

**Discussion:** G-ratio and MTsat changes in NAWM over one year indicate subtle myelin loss in early RRMS, which were not apparent with NAWM MTR or in WBV. Increases in NAWM and WML NODDI ICVF were not anticipated, and raise the possibility of axonal swelling or morphological change. Increases in WML MTsat may reflect myelin repair. Changes in NODDI ISOVF are more likely to reflect alterations in water content. Competing MTsat and ICVF changes may account for the absence of g-ratio change in WML. Longitudinal changes in microstructural measures are significant at a group level, however detection in individual patients in early RRMS is limited by technique reproducibility.

**Conclusion:** MTsat and g-ratio are more sensitive than MTR to early pathological changes in RRMS, but complex dependence of g-ratio on NODDI parameters limit the interpretation of aggregate measures in isolation. Improvements in technique reproducibility and validation of MRI biophysical models across a range of pathological tissue states are needed.

## 1. Introduction

### 1.1 The need for longitudinal microstructural markers in multiple sclerosis (MS)

MS is a chronic, immune-mediated neurodegenerative disease of the central nervous system (CNS) with heterogeneous symptomology, including motor impairment, fatigue, visual disturbances, and variable disease course (Brownlee et al., 2017; Filippi et al., 2018; Gelfand, 2014). Inflammation-associated demyelination is believed to result in axonal degeneration that ultimately causes disability (Bitsch et al., 2000; Frischer et al., 2009; Lassmann, 2018). Relapsing-remitting MS (RRMS) is characterised by clinical relapses interspersed with periods of remission, and it is difficult to predict at disease onset which patients will subsequently experience a more aggressive disease course (Scalfari et al., 2010).

Magnetic resonance imaging (MRI) is widely used in clinical practice for diagnosis and tracking MS activity over time. Conventional structural MRI sequences demonstrate white matter lesions (WML) although provide limited specificity for charactering damage within these, and are insensitive to abnormalities such as decreased fibre density (Kutzelnigg et al., 2005) and subtle demyelination (Allen and McKeown, 1979) in normal-appearing white matter (NAWM). Longitudinal measurements of brain volume provide downstream indicators of neurodegeneration but lack specificity for these disease-relevant processes and are confounded by other factors (Wattjes et al., 2021). Candidate *in vivo* microstructural MRI markers, that are sensitive and specific to early changes in myelin and axonal integrity, are thus needed for tracking and predicting disease trajectory, and evaluating response to disease-modifying therapies (DMTs), putative remyelinating and neuroprotective treatments.

### 1.2 Magnetisation transfer imaging (MTI)

MTI derives signal indirectly from protons ‘bound’ to macromolecules, which tend to be myelin-associated in the CNS. The T2 of ‘bound’ protons is shorter than normal echo times (∼10μs), and thus typically not MRI-visible. The derived magnetisation transfer ratio (MTR) has been extensively applied in cohorts of RRMS (York et al., 2022), but its use as a surrogate endpoint in large, multi-centre clinical trials is limited by lack of sensitivity to subtle demyelination in NAWM (Bonnier et al., 2014; Cercignani et al., 2009), poor reproducibility and sensitivity to variation in scanning acquisition parameters (Helms et al., 2010a; Tofts et al., 2006).

Measures such as magnetisation transfer saturation (MTsat) (Helms et al., 2008a) and inhomogeneous MTR (ihMTR) (Varma et al., 2015) present clinically feasible alternatives to time-consuming fully quantitative MTI approaches while overcoming some of the limitations of MTR. MTsat inherently corrects for T1 relaxation and B1 inhomogeneities (Helms et al., 2008a). In MS, MTsat is lower in NAWM in MS than healthy control white matter (Lommers et al., 2019), and lower NAWM and WML MTsat in the brain and cervical spinal cord are associated with worse clinical disability (Lema et al., 2017). MTsat may therefore be more specific to changes in myelin integrity than MTR, but has not previously been studied longitudinally in recently diagnosed RRMS.

### 1.3 Diffusion-weighted imaging (dMRI)

dMRI is sensitive to neuroaxonal structures, but relatively insensitive to myelin. In the presence of highly structured white matter tracts, the measured water diffusion is anisotropic but becomes increasingly isotropic with neuronal degeneration. Glial cell infiltration (Yi et al., 2019) and crossing fibres may, however, complicate biological interpretation (Jones et al., 2013).

Modelling the dMRI signal from multi-shell acquisition protocols may help to resolve structural uncertainty. Neurite Orientation Dispersion and Density Imaging (NODDI) (Zhang et al., 2012), for example, considers the diffusion signal as isotropic (ISOVF), restricted (ICVF) and hindered diffusion volume fractions, plus an orientation dispersion index (ODI) (Zhang et al., 2012). The NODDI model is based on a number of assumptions (Zhang et al., 2012), including a fixed intrinsic diffusivity rate, but has previously been applied in studies of MS (Alotaibi et al., 2021; Schneider et al., 2017). ICVF may be useful as a marker of neurite (axon and dendrite) density and is reduced in WML in RRMS compared with healthy control white matter, while results for NAWM are mixed (Collorone et al., 2020; Hagiwara et al., 2019; Rahmanzadeh et al., 2021). NODDI metrics may provide useful early markers of neuroaxonal degeneration, however NODDI changes with time in RRMS are largely unexplored (Alotaibi et al., 2021).

### 1.4 Aggregate MRI g-ratio

Measures which combine microstructural MRI methods, such as the MRI aggregate g-ratio, may also better capture the net effects of disease and/or treatment than an individual imaging biomarker alone (Stikov et al., 2015; York et al., 2021). The g-ratio is a measure of myelin thickness, defined as the ratio of the diameter of the neuronal axon to the diameter of the myelinated axon (Rushton, 1951). Originally a neuropathological measure, a theoretical optimal g-ratio of 0.6 for maximum neuronal transduction was proposed (Rushton, 1951), although later work suggests a higher value (0.72-0.81 in the CNS) is more realistic (Chomiak and Hu, 2009). Abnormally high g-ratios are indicative of myelin disruption (Ellerbrock and Mohammadi, 2018; Kamagata et al., 2019).

G-ratio parametric maps may be derived by combining MTsat and NODDI data on a voxel-by-voxel basis (Campbell et al., 2018; Hori et al., 2018; Kamagata et al., 2019; Stikov et al., 2015; York et al., 2021). Although dependent on a number of prior assumptions that have been extensively reviewed elsewhere (Campbell et al., 2018; Mohammadi and Callaghan, 2021), the MRI g-ratio has been validated against *ex vivo* electron microscopy in the macaque (Stikov et al., 2015). In RRMS, increased g-ratios have shown an association with elevated plasma neurofilament, a blood marker of active axonal damage (York et al., 2021), and g-ratio structural connectome disruption has been related to disease severity (Kamagata et al., 2019). Again, the limited published studies of g-ratios in MS have been cross-sectional, rather than measuring neurodegeneration with time.

### 1.5 Rationale and Aims

This study aims to evaluate *in vivo* markers of microstructural integrity for early disease stratification and as surrogate endpoints for future therapeutic trials. To this end, the sensitivity of MTR, MTsat, NODDI and g-ratio measures for detecting pathological change with time were compared in WML and NAWM in recently diagnosed RRMS; and with whole brain atrophy, an established general marker of neurodegeneration. To establish the applicability of these parametric changes to individual patients, the magnitude of these changes was compared with technique test-retest agreement in a group of health controls.

## 2. Materials and Methods

### 2.1 Participants

#### 2.1.1 Patients with relapsing-remitting multiple sclerosis

Seventy-nine people with recently diagnosed RRMS were recruited to a longitudinal single-centre sub-study of FutureMS at the Anne Rowling Regenerative Neurology Centre (Edinburgh, Scotland). FutureMS (Kearns et al., 2022; Meijboom et al., 2022) is a multicentre, prospective, longitudinal cohort study of 440 people with RRMS, who were diagnosed within the previous six months, according to 2010 McDonald criteria (Polman et al., 2011). Individuals with MS were required to be over 18 years of age and the baseline assessment was prior to initiation of any DMT. Visits at baseline (M0) and one-year follow-up (M12) included MRI and clinical assessment.

#### 2.1.2 Healthy controls

Twelve healthy volunteers were additionally imaged with the same MRI protocol, which was repeated within two weeks to determine test-retest agreement.

### 2.2 Ethical Approval

Approval for the sub-study was obtained from the local Research Ethics Committee (reference REC 15/SS/0233). The study conformed to the Declaration of Helsinki 2000 (amendments in 2002 and 2004) and Good Clinical Practice ICH guidelines. All participants provided written informed consent.

### 2.3 MRI acquisition

All images were acquired on a 3.0 T Prisma MRI system (Siemens, Erlangen, DE) at the Edinburgh Imaging Facility (Royal Infirmary of Edinburgh) with a 32 channel head coil.

Structural images included a 3D T1-weighted MPRAGE, 2D and 3D FLAIR, and 2D T2-weighted dual echo sequences (see Table 1 for full MRI acquisition parameters). MTI consisted of three consecutive 3D gradient-echo FLASH sequences: two proton density images with and without a Gaussian off-resonance MT saturation pulse (MT_on_ and MT_off_, respectively), and an additional T1-weighted image. Multi-shell diffusion-weighted 2D spin-echo echo-planar imaging was also performed with 151 diffusion directions and three reverse phase encoding volumes.

**Table 1:**
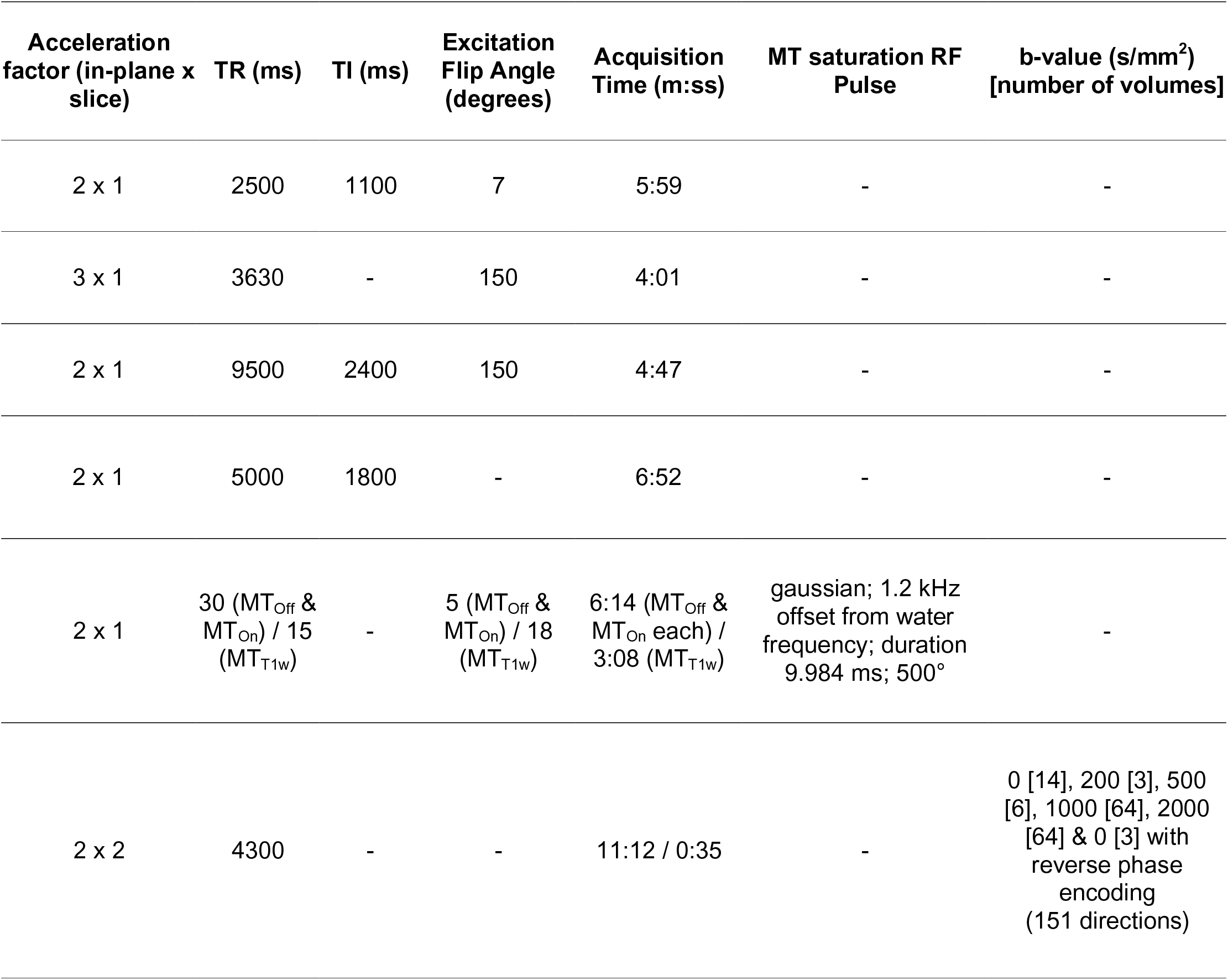
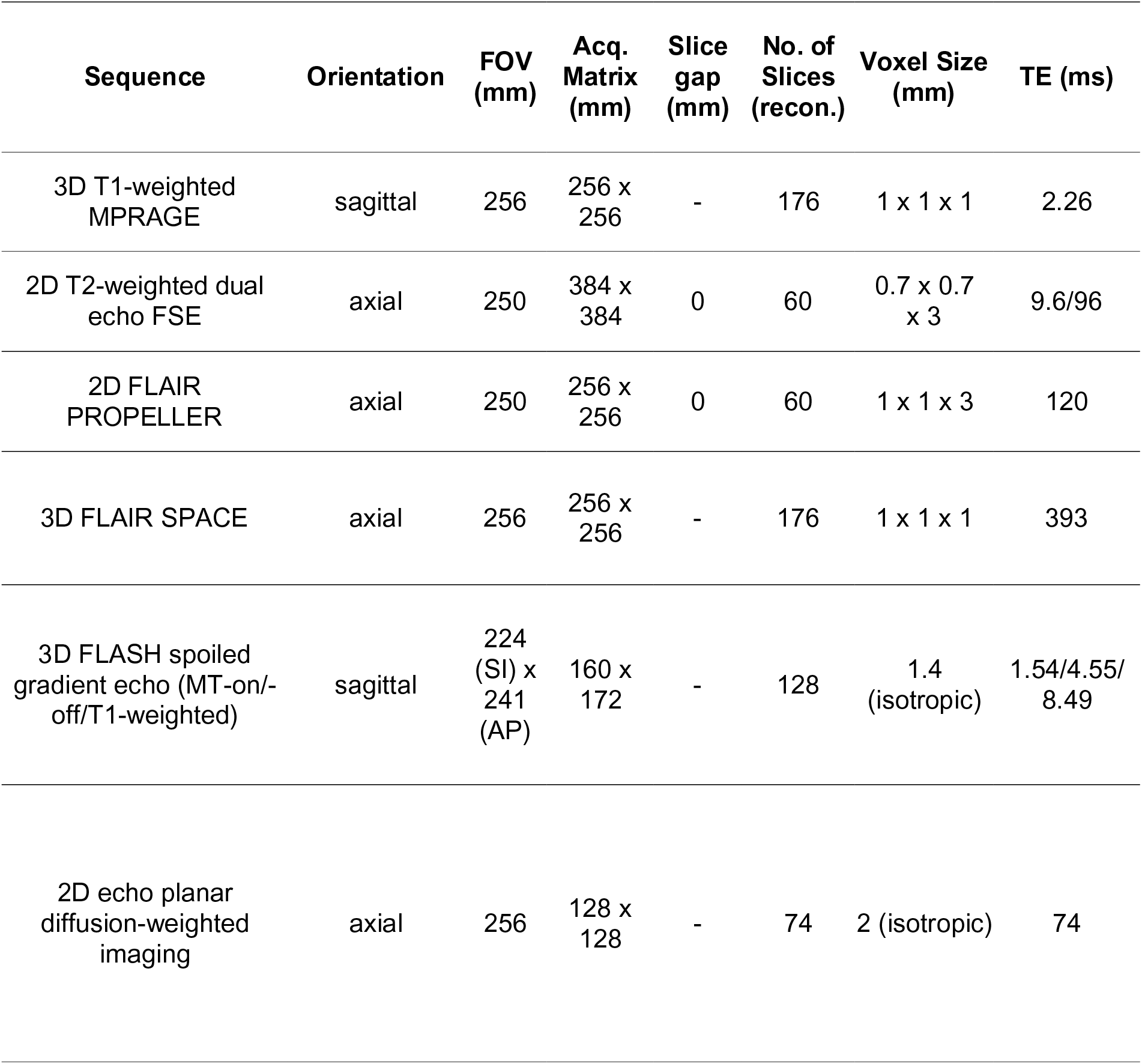
MRI acquisition parameters for structural, magnetisation transfer (MT) and diffusion-weighted imaging (dMRI). Acq. matrix: acquisition matrix; FOV: field of view; recon.: reconstructed; RF: radiofrequency; TE: echo time; TI: inversion time; TR: repetition time.

| Acceleration factor (in-plane x slice) | TR (ms) | TI (ms) | Excitation Flip Angle (degrees) | Acquisition Time (m:ss) | MT saturation RF Pulse | b-value (s/mm<sup>2</sup>) [number of volumes] |
| --- | --- | --- | --- | --- | --- | --- |
| 2 x 1 | 2500 | 1100 | 7 | 5:59 | - | - |
| 3 x 1 | 3630 | - | 150 | 4:01 | - | - |
| 2 x 1 | 9500 | 2400 | 150 | 4:47 | - | - |
| 2 x 1 | 5000 | 1800 | - | 6:52 | - | - |
| 2 x 1 | 30 (MT<sub>Off</sub> & MT<sub>On</sub>) / 15 (MT<sub>T1w</sub>) | - | 5 (MT<sub>Off</sub> & MT<sub>On</sub>) / 18 (MT<sub>T1w</sub>) | 6:14 (MT<sub>Off</sub> & MT<sub>On</sub> each) / 3:08 (MT<sub>T1w</sub>) | gaussian; 1.2 kHz offset from water frequency; duration 9.984 ms; 500° | - |
| 2 x 2 | 4300 | - | - | 11:12 / 0:35 | - | 0 [14], 200 [3], 500 [6], 1000 [64], 2000 [64] & 0 [3] with reverse phase encoding (151 directions) |

| Sequence | Orientation | FOV (mm) | Acq. Matrix (mm) | Slice gap (mm) | No. of Slices (recon.) | Voxel Size (mm) | TE (ms) |
| --- | --- | --- | --- | --- | --- | --- | --- |
| 3D T1-weighted MPRAGE | sagittal | 256 | 256 x 256 | - | 176 | 1 x 1 x 1 | 2.26 |
| 2D T2-weighted dual echo FSE | axial | 250 | 384 x 384 | 0 | 60 | 0.7 x 0.7 x 3 | 9.6/96 |
| 2D FLAIR PROPELLER | axial | 250 | 256 x 256 | 0 | 60 | 1 x 1 x 3 | 120 |
| 3D FLAIR SPACE | axial | 256 | 256 x 256 | - | 176 | 1 x 1 x 1 | 393 |
| 3D FLASH spoiled gradient echo (MT-on/-off/T1-weighted) | sagittal | 224 (SI) x 241 (AP) | 160 x 172 | - | 128 | 1.4 (isotropic) | 1.54/4.55/ 8.49 |
| 2D echo planar diffusion-weighted imaging | axial | 256 | 128 x 128 | - | 74 | 2 (isotropic) | 74 |

### 2.4 MRI Processing

#### 2.4.1 Brain tissue segmentation

Structural MRI data processing is described in detail elsewhere.^38^ Briefly, all images were first converted from DICOM to NIfTI format (dcm2niix v1.0.20190410 (Li et al., 2016)). For people with RRMS, WML were defined as hyperintensities on T2 FLAIR at M0 and segmented automatically using an in-house thresholding approach (Meijboom et al., 2022), with manual correction where necessary (ITK-SNAP v3.6, http://www.itksnap.org). At follow-up, baseline WML masks were registered to follow-up FLAIR images and manually edited for changes.

Structural T1-weighted MPRAGE images were skull-stripped (FSL v6.0.1) and brain tissue segmentation (whole brain and NAWM) was carried out with FreeSurfer (v6.0, https://surfer.nmr.mgh.harvard.edu/) at each time point, followed by FreeSurfer’s longitudinal processing stream. Visual quality assurance checks and correction when needed were performed. Whole-brain volume (WBV) was measured using fslstats (FSL v6.0.1) and corrected for intracranial volume.

#### 2.4.2 MTI parametric maps

Using an in-house MATLAB script (R2018b, software available: https://doi.org/10.7488/ds/2965, requires SPM12 and FSL functions) (York, 2020), echoes were summed together to increase the signal-to-noise ratio (SNR) (Helms and Hagberg, 2009) for each MT image (MT_on_, MT_off_, and MT_T1w_). MT_on_ and MT_T1w_ images were registered to the MT_off_ image with a rigid-body transformation (6 degrees of freedom, FSL FLIRT (Jenkinson et al., 2002; Jenkinson and Smith, 2001)). Parametric MTsat maps (Figure 1) were calculated from MT images (MT_on_, MT_off_, and MT_T1w_), as detailed previously, including correction of approximated T1 (Helms et al., 2008b; Helms et al., 2010b; Helms et al., 2008a). MTR maps (Figure 1) were calculated as MTR = 100 x (MT_off_-MT_on_/MT_off_).

**Figure 1:**
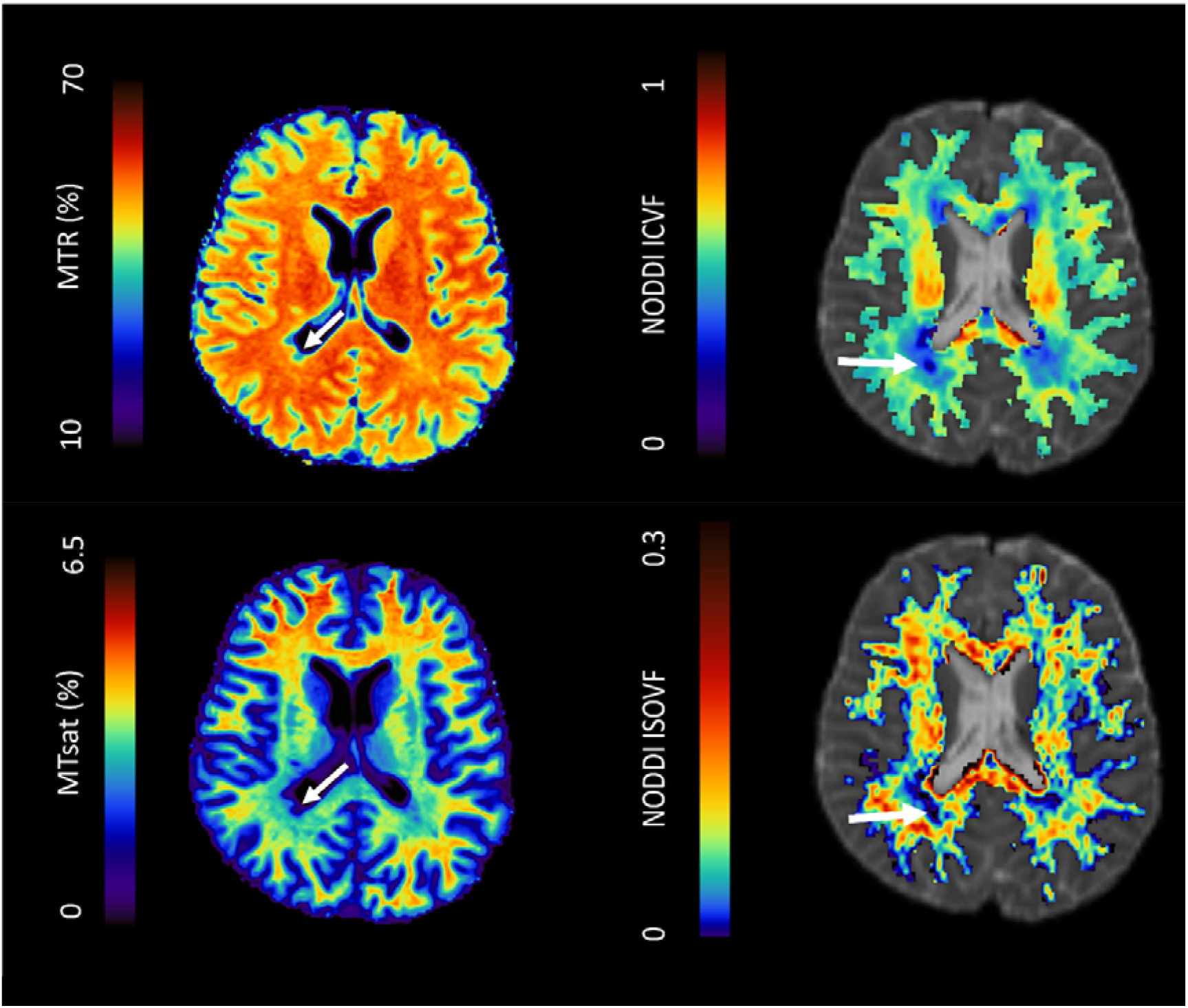
Example parametric maps for a person with recently diagnosed relapsing-remitting multiple sclerosis: magnetisation transfer ratio (MTR, top left), magnetisation transfer saturation (MTsat, bottom left), Neurite Orientation Dispersion and Density Imaging (NODDI) intracellular volume fraction (ICVF, top right) and isotropic volume fraction (ISOVF, bottom right). Arrows: white matter lesion.

#### 2.4.3 NODDI parametric maps

dMRI processing included brain extraction and removal of bulk motion and eddy-current-induced distortions with FSL (v6.0.1). All dMRI volumes were registered to the first b0 diffusion volume before processing with the NODDI toolbox (Zhang et al., 2012) (v1.0, mig.cs.ucl.ac.uk, MATLAB R2016b) to produce ICVF and ISOVF parametric maps (Figure 1) (Meijboom et al., 2022).

#### 2.4.4 G-ratio parametric maps

MTsat maps were registered to dMRI b0 reference volumes before calculating g-ratio maps (FSL epi_reg). Creation of g-ratio maps (Figure 2) followed methodology detailed previously (York et al., 2021; Zhang et al., 2012), using equation from Stikov et al., (2015):

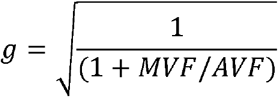

**Figure 2:**
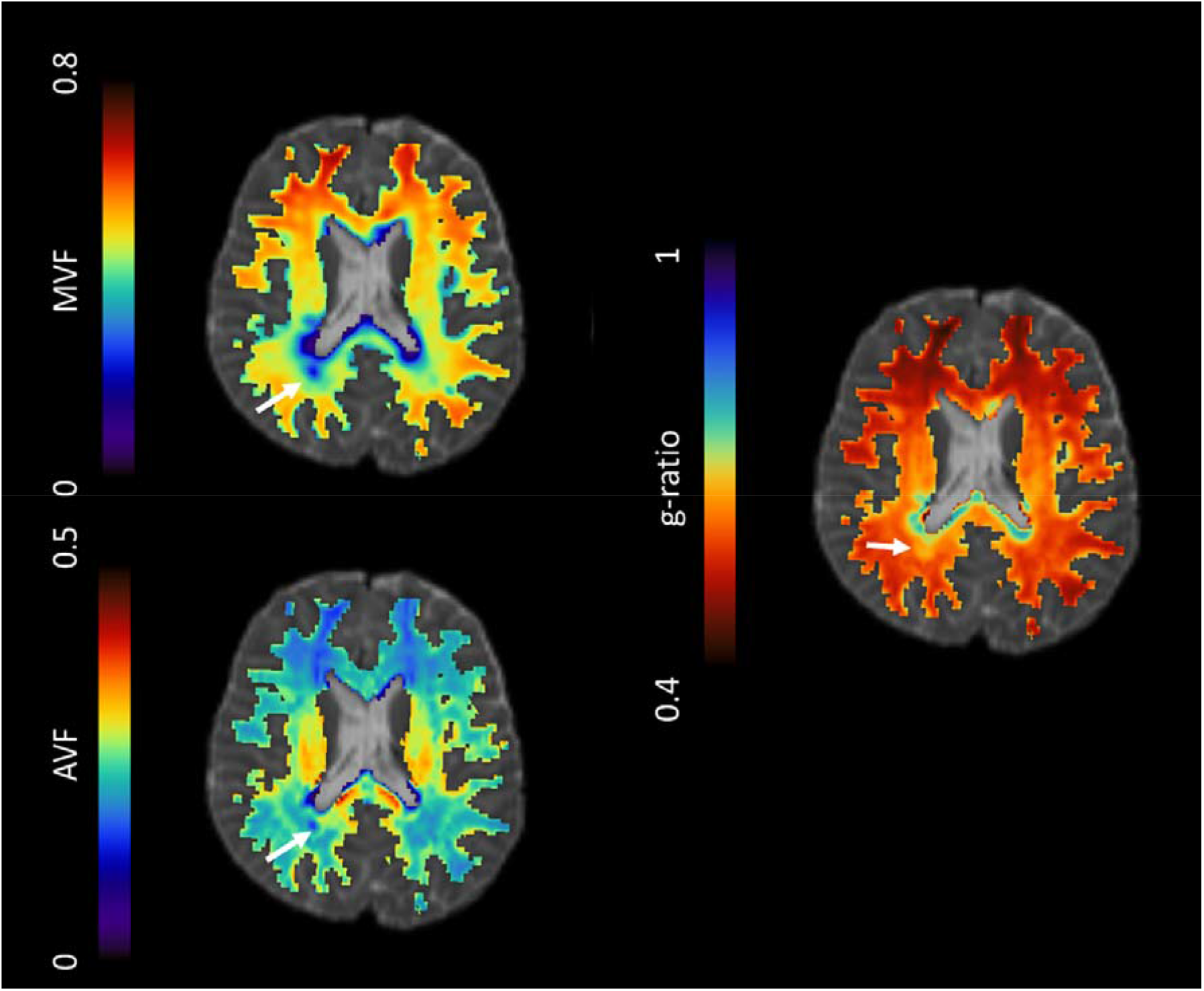
Example myelin volume fraction (MVF, top left), axonal volume fraction (AVF, bottom left) and g-ratio parametric maps (right) for a patient with recently diagnosed relapsing-remitting multiple sclerosis. Arrows: white matter lesion.

MVF is the myelin volume fraction derived from linearly-scaled MTsat maps (Campbell et al., 2018); AVF is the axonal volume fraction derived from NODDI dMRI data, calibrated in healthy control subjects (York et al., 2021). The cerebellum was not included in dMRI and g-ratio analyses due to technical inaccuracies.

#### 2.4.5 Mask-to-map registration

For each time-point, tissue segmentations were registered to either the MT_off_ image (FSL FLIRT (Jenkinson et al., 2002; Jenkinson and Smith, 2001)) for MT maps or the first b0 volume of diffusion data for g-ratio maps. To minimise partial volume effects, erosion by one voxel was applied to NAWM masks.

### 2.5 Statistical and Graphical Analyses

All statistical analyses were performed in RStudio (v1.4.1717, R v3.6.1).

#### 2.5.1 Descriptive statistics

Tissue masks were applied to parametric maps (in-house code with *RNifti* package v1.3.0) to output summary statistics (e.g. mean, median).

#### 2.5.2 Test-retest agreement of microstructural metrics in healthy white matter

Bland-Altman plots (Martin Bland and Altman, 1986) (*BlandAltmanLeh* R package) were used to assess test-retest agreement from healthy control data. Here, the difference in mean values between time points is plotted against the average value over time points for each subject and each microstructural metric. The limits of agreement show the range within which 95% of subjects would be expected to fall if measures were repeated in healthy control white matter, and thus establishes reference levels for pathological change. Sign tests were used to determine whether the difference between time points was significantly different from zero (significance level, α=0.05).

#### 2.5.3 Longitudinal change

Longitudinal changes in MTsat, MTR, ISOVF, ICVF and g-ratio across NAWM and WML in RRMS patients were first assessed using paired t-tests (α=0.05). When significant, follow-up linear mixed modelling (maximum likelihood) was performed to account for potential confounding variables (age, sex, lesion load [as a percentage of intracranial volume] and initiation of DMTs; *R* packages *lme4* and *lmerTest* (Bates et al., 2015; Kuznetsova et al., 2017)). Whole brain atrophy was also examined with a linear mixed model. Interaction terms were included in models where appropriate.

Post-hoc false discovery rate (FDR) correction for multiple comparisons was performed for linear mixed models. Goodness-of-fit was assessed with Nakagawa’s marginal R^2^ for mixed models (Nakagawa et al., 2017) (R package *performance* (Lüdecke et al., 2021)).

The relationship between longitudinal change across microstructural metrics, and with whole brain atrophy was examined with Pearson’s correlation coefficients. *Post-hoc* comparisons of longitudinal changes between patients with and without new lesions at M12, identified by an experienced neuroradiologist, was performed with Welch’s t-tests.

#### 2.5.4 Comparison with healthy control test-retest agreement

To compare longitudinal change over one year with test-retest agreement, the mean difference and limits of agreement from NAWM Bland-Altman plots from test-retest data acquired in healthy controls were superimposed on boxplots of longitudinal change.

#### 2.5.5 Simulating pathological change

To understand how individual changes in myelin and/or axonal volume fractions would affect the g-ratio, in a *post hoc* simulation, biologically realistic parametric values for MTsat and NODDI measures were substituted into equations to calculate MVF, AVF and g-ratio, and plotted in R (*ggplot* package).

## 3. Results

### 3.1 Demographics

#### 3.1.1 Healthy Controls

Data from eleven healthy controls (7 females, mean age 44 years [range 27-58 years]) contributed to the final analysis. One healthy control was excluded due to an unexpected incidental imaging finding. Due to dMRI processing errors at the second time-point, two controls were excluded from NODDI and g-ratio test-retest analyses.

#### 3.1.2 People with RRMS

Seventy-nine patients with RRMS underwent imaging at M0. Two were excluded from all analyses as more than six months had passed between diagnosis and imaging. Fifteen datasets were excluded from longitudinal analyses due to poor FreeSurfer tissue segmentation (n=5), missing M12 MRI data (n=8), and failure to complete the full MRI protocol at M0 (n=1) or M12 (n=1). In addition, dMRI processing failed for two subjects. Longitudinal MTI data were hence available for 62 patients, and dMRI data were available for 60 patients (see Table 2 for demographics).

**Table 2:** Demographics for people with RRMS with complete data at M0 (baseline) and follow-up (M12). DMT: disease-modifying therapy; EDSS: Expanded Disability Status Scale score; ICV: intracranial volume; WML: white matter lesions

|  | Magnetisation transfer imaging | NODDI ISO/ICVF & g-ratio |
| --- | --- | --- |
| n (F:M) | 62 (48:14) | 60 (46:14) |
| Mean age [range] in years at M0 | 37.6 [21.7 to 67.3] | 37.8 [22.3 to 67.3] |
| Median EDSS at M0 [range] | 2 [0 to 6] | 2 [0 to 6] |
| Median EDSS at M12 [range] | 2.5 [0 to 6.5] | 2.75 [0 to 6.5] |
| Number of patients with DMT initiated by M12 | 38 (61.3%) | 36 (60.0%) |
| Mean number of days between diagnosis and M0 [range] | 68 [7 to 171] | 69 [8 to 171] |
| Median time since physician-reported first symptom onset in years [range] | 3.6 [0.2 to 33.2]<br>(2 missing) | 3.55 [0.2 to 33.2]<br>(2 missing) |
| Median WML volume (as % of ICV) at M0 [range] | 0.572<br>[0.041 to 2.628] | 0.572<br>[0.041 to 2.628] |
| Median change in WML volume over 1 year (as % of ICV, abs. diff) [range] | 0.165<br>[0.028 to 0.592] | 0.165<br>[0.028 to 0.592] |

### 3.2 Test-retest agreement

Descriptive statistics and Bland-Altman limits of agreement for healthy white matter are reported in Table 3. Sign tests and Bland-Altman plots (Appendix Figs. A.1-2) show that the mean difference did not differ from zero between time points for any microstructural metric (all p>0.05, Table 3).

**Table 3:** Mean (standard deviation) MTI, NODDI and g-ratio values in healthy control white matter (n=11, except where indicated *n=9). P-values are given for two-sided sign tests for matched pairs. ^$^excludes cerebellum

|  | Time point 1 | Time point 2 | Bland-Altman |  | p-value (uncorrected) |
| --- | --- | --- | --- | --- | --- |
|  |  |  | mean difference | limits of agreement |  |
| **MTsat (%)** | 3.74 (0.13) | 3.78 (0.11) | 0.04 (0.09) | ±0.186 | 0.549 |
| **MTR (%)** | 54.51 (0.66) | 54.25 (0.70) | -0.26 (0.68) | ±1.341 | 1.00 |
| **NODDI**<br>**ISOVF<sup>§</sup>** | 0.086<br>(0.008) | 0.085<br>(0.009)* | -0.001<br>(0.003)* | ±0.0057* | 1.00 |
| **NODDI**<br>**ICVF<sup>§</sup>** | 0.605<br>(0.022) | 0.603<br>(0.020)* | -0.002<br>(0.003)* | ±0.0063* | 0.508 |
| **g-ratio<sup>§</sup>** | 0.581<br>(0.015) | 0.576<br>(0.012)* | -0.005<br>(0.012)* | ±0.024* | 0.508 |

### 3.3 Longitudinal microstructural change in recently diagnosed RRMS

#### 3.3.1 Longitudinal change in ‘normal-appearing’ white matter

In NAWM, paired t-tests show a significant decrease in MTsat and a significant increase in NODDI ICVF and g-ratio over one year (Table 4). Group mean changes over time (Table 4) were lower than the limits of agreement established from healthy control data (Table 3), although a small number of subjects exceeded these limits (Appendix Fig. A.4). MTR and NODDI ISOVF did not change significantly over one year.

**Table 4:** Descriptive statistics and paired t-tests for MTI (n=62), g-ratio and NODDI (n=60) data in ‘normal-appearing’ white matter. MTsat: magnetisation transfer saturation; MTR: magnetisation transfer ratio; ICVF: intracellular volume fraction; ISOVF: isotropic volume fraction; SD: standard deviation; M0: baseline; M12: one year follow-up. *excludes cerebellum for g-ratio and NODDI metrics

| **'Normal-appearing' white matter\*** |  |  |  |  |  |
| --- | --- | --- | --- | --- | --- |
|  | **Mean [range]** |  | **mean**<br>**diff. (SD)** | **paired t-test** |  |
|  | **M0** | **M12** |  | **t-value** | **p-value**<br>**(uncorrected)** |
| **MTsat** | 3.80 | 3.77 | -0.03 | -2.19 | 0.033 |
| **(%)** | [3.43 to 4.07] | [3.39 to 4.25] | [0.12] |  |  |
| **MTR** | 54.25 | 54.24 | -0.01 | -0.079 | 0.94 |
| **(%)** | [50.80 to 56.28] | [52.02 to 56.39] | [0.82] |  |  |
| **g-ratio** | 0.57 | 0.574 | 0.004 | 2.60 | 0.012 |
|  | [0.538 to 0.609] | [0.523 to 0.625] | [0.012] |  |  |
| **NODDI** | 0.075 | 0.074 | 0 | -0.43 | 0.67 |
| **ISOVF** | [0.054 to 0.097] | [0.044 to 0.096] | [0.007] |  |  |
| **NODDI** | 0.577 | 0.579 | 0.002 | 2.29 | 0.025 |
| **ICVF** | [0.504 to 0.640] | [0.499 to 0.639] | [0.007] |  |  |

After controlling for age, lesion load, sex, initiation of DMTs and interaction terms, linear mixed models showed that the effect of time on NAWM g-ratio (β = 0.005, *t*(75.91) = 3.08, FDR-corrected *p* = 0.006, Appendix Table A.1), NODDI ICVF (β = 0.003, *t*(90.89) = 3.51, FDR-corrected *p* = 0.002, Appendix Table A.2) and MTsat remained significant (β = -0.040, *t*(79.26) = -2.60, FDR-corrected *p* = 0.018, Appendix Table A.3).

In NAWM, one year change in g-ratio was strongly associated with change in MTsat (Pearson’s R^2^ = 0.98, *p*<0.001, Appendix Fig. A.5). Correlations between change in NODDI metrics and g-ratio were weaker but statistically significant (Pearson’s R^2^ = 0.18 and 0.11, *p*<0.001 and *p* = 0.011 for NODDI ICVF and ISOVF, respectively), as were associations between longitudinal change in MTsat and NODDI metrics (Pearson’s R^2^ = 0.11 and *p* = 0.009 for both ICVF and ISOVF).

#### 3.3.2 Longitudinal change in white matter lesions

In WML, paired t-tests (Table 5) and linear mixed models reveal significant longitudinal increases in MTsat (β = 0.059, *t*(82.58) = 3.65, FDR-corrected *p* = 0.002, Appendix Table A.4), NODDI ICVF (β = 0.017, *t*(80.64) = 6.95, FDR-corrected *p* = 0.002, Table A.5) and ISOVF (β = 0.011, *t*(74.85) = 5.56, FDR-corrected *p* = 0.002,

**Table 5:**
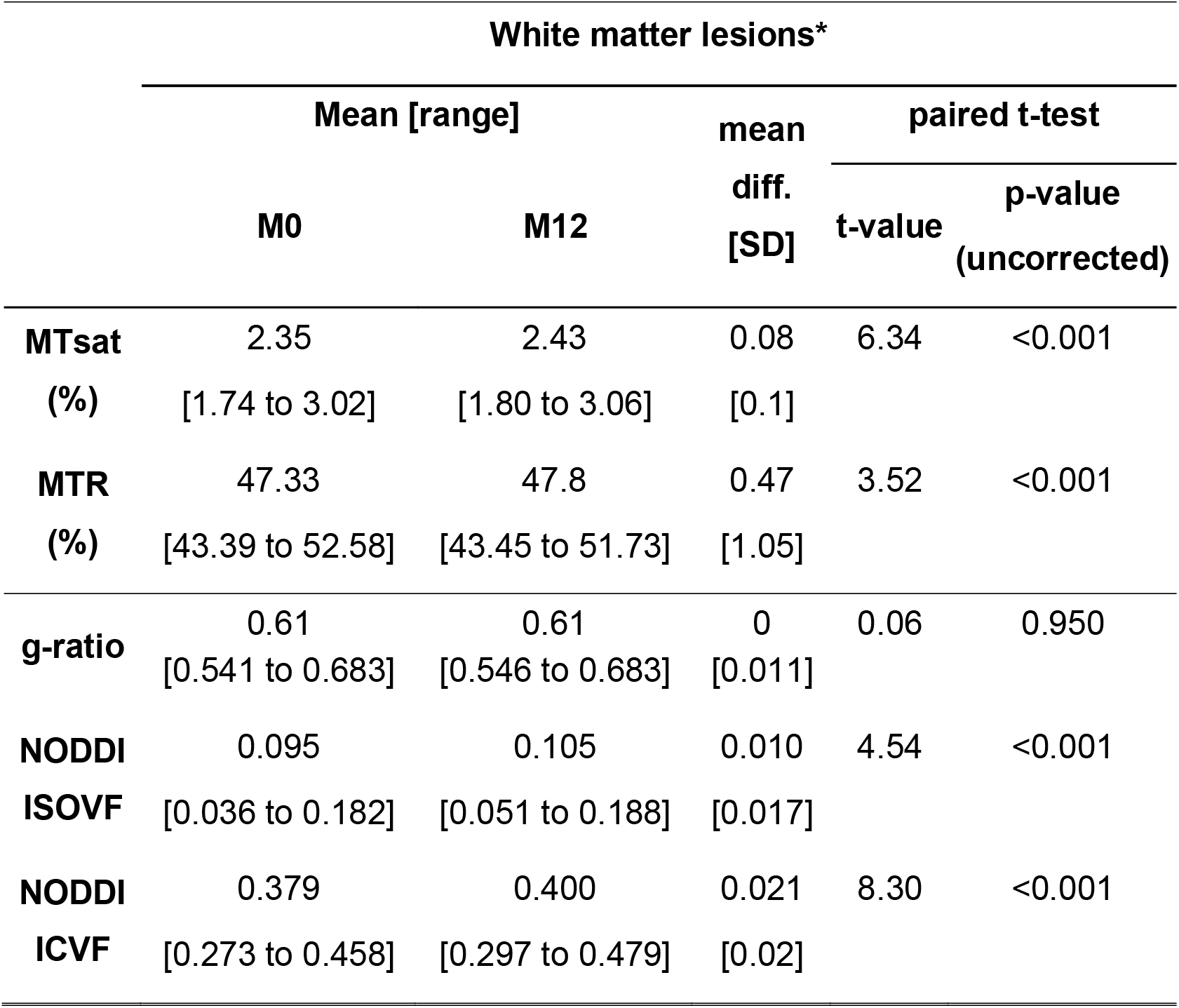
Descriptive statistics and paired t-tests for MTI (n=62), g-ratio and NODDI (n=60) data in T2 FLAIR white matter lesions. MTsat: magnetisation transfer saturation; MTR: magnetisation transfer ratio; ICVF: intracellular volume fraction; ISOVF: isotropic volume fraction; SD: standard deviation; M0: baseline; M12: one year follow-up. *excludes cerebellum for g-ratio and NODDI metrics

Table A.6). The change in MTR was not significant after adjusting for confounding factors (β = 0.24, *t*(79.48) = 1.60, *p* = 0.113, Table A.7) and there was no change in g-ratio over the same time period (see Table 6 for summary of results).

**Table 6:** Summary of linear mixed model results. MTsat: magnetisation transfer saturation; MTR: magnetisation transfer ratio; NODDI: neurite orientation dispersion and density index; ISOVF: isotropic volume fraction; ICVF: intraneurite volume fraction; NAWM: normal-appearing white matter; WML: white matter lesions; ^$^cerebellum excluded; *significant after False Detection Rate correction for multiple comparisons.

|  | Longitudinal Change |  |
| --- | --- | --- |
|  | NAWM | WML |
| **MTsat** | ↓ * | ↑ * |
| **MTR** | - | - |
| **g-ratio<sup>\$</sup>** | ↑ * | - |
| **NODDI ISOVF<sup>\$</sup>** | - | ↑ * |
| **NODDI ICVF<sup>\$</sup>** | ↑ * | ↑ * |

Group mean changes in MTsat and MTR lay within limits of agreement established in healthy control white matter (Table 3). Group-wise increases in NODDI ISOVF and ICVF were greater than the limits of agreement and a large number of individual subjects exceeded both positive and negative limits of agreement for both metrics (Appendix Fig. A.4).

In WML, associations between longitudinal change in g-ratio and other metrics were weak but significant (Pearson’s R^2^ = 0.16, 0.17 and 0.11, *p* = 0.002, <0.001 and 0.011 for MTsat, NODDI ICVF and ISOVF, respectively, Appendix Fig. A.6). Change in MTsat within WML was moderately associated with NODDI ICVF (Pearson’s R^2^ = 0.29, p<0.001) and weakly negatively associated with NODDI ISOVF (Pearson’s R^2^ = 0.16, p=0.002).

### 3.4 Whole brain atrophy

After adjusting for covariates, there was no signifcant decrease in WBV over one year (Appendix Table A.8) and no relationship was found between whole brain atrophy and any of the microstructural measures that showed significant change over time (Appendix Fig. A.7).

### 3.5 Presence of new lesions

There were no significant differences in mean change over time between patients who did or did not have new lesions at M12 (FDR-corrected p>0.05 for all metrics).

### 3.6 Simulating pathological change

Simulated data points (Appendix Fig. A.3) show the competing effects of changes in MTsat and NODDI metrics on g-ratio. As MTsat decreases, g-ratio increases, when NODDI metrics are held constant. An increase in ICVF, however, may also increase the g-ratio while an increase in ISOVF lowers the g-ratio.

## 4. Discussion

### 4.1 Summary of Results

In this study, changes in microstructural MRI measures in WML and NAWM were assessed over the year following diagnosis of RRMS. In NAWM, there was a decrease in MTsat and an increase in g-ratio and NODDI ICVF, but no change in MTR or NODDI ISOVF (see summary Table 6). In WML, a longitudinal increase in MTsat, NODDI ICVF and NODDI ISOVF but no change in g-ratio or MTR was demonstrated. Despite significant group-wise changes, the majority of individual patients remain within test-retest limits of agreement in healthy white matter, with the exception of NODDI measures in cerebral WML. No significant whole brain atrophy was detectable, nor did atrophy correlate with any microstructural measure.

Simulated data indicate that biological interpretation of longitudinal change in g-ratio is complicated due to its inverse relationship with MTsat, positive dependence on NODDI ICVF and a negative dependence on NODDI ISOVF.

### 4.2 Longitudinal changes in NAWM

#### 4.2.1 MTI

The longitudinal decline in MTsat observed in NAWM suggests that subtle loss of myelin integrity occurs in recently diagnosed RRMS, which cannot be seen on conventional T2-weighted FLAIR. Longitudinal MTsat data in RRMS have not previously been reported, although MTsat is lower in NAWM in MS than healthy control white matter (Lommers et al., 2019). The reduction in MTsat in NAWM over one year in our study was small in comparison to variance in test-retest healthy control white matter. This suggests that, although there may be a weak group-wise longitudinal change in NAWM MTsat, measurement error may limit application on an individual patient level (e.g. for clinical decision-making). Patients in our study were recruited shortly after diagnosis, and resolving effects associated with the acute inflammatory episode that prompted diagnosis at baseline are potential confounds; heterogeneous demyelination and myelin repair across NAWM could therefore also contribute to the weak effect over a relatively short period early in disease.

Nevertheless, results indicate that MTsat is more sensitive to early RRMS pathology in NAWM than MTR, which shows no detectable change. While MTsat and MTR are both sensitive to protons ‘bound’ to macromolecules within the lipid bilayers of myelin, MTR signal also depends non-linearly on T1 recovery effects and B1 inhomogeneities, which are effectively corrected for in MTsat (Helms et al., 2008a). T1 prolongation, which accompanies myelin damage (Al-Radaideh et al., 2015), may systematically affect MTR in such a way as to render it less sensitive to demyelination than MTsat (Helms et al., 2008a). Previous case-control studies show that MTR in NAWM is typically only 1.25 percent units lower than control white matter and the longitudinal decline in NAWM MTR has been estimated at 0.1% per year (Davies et al., 2005), reiterating the subtlety of NAWM changes (York et al., 2022). Longer follow-up may therefore be required in order to detect changes in NAWM MTR in early RRMS.

#### 4.2.2 NODDI

An unexpected increase in NODDI ICVF was seen within NAWM over one year. NODDI ICVF (the ‘restricted’ diffusion signal fraction) is typically lower in RRMS compared with healthy controls (Alotaibi et al., 2021; Collorone et al., 2020; Johnson et al., 2021a), although this is not a universal finding (Kato et al., 2022). The established literature on longitudinal NODDI measurements is sparse, but decreases in NAWM ICVF over time have previously been noted (Sacco et al., 2020). A ‘borderline’ significant increase in mean fractional anisotropy (FA) within ‘normal-appearing’ brain tissue over two years has been reported elsewhere (Rashid et al., 2008), but the annualised rate of change did not differ from healthy control subjects.

The biological mechanism underlying the observed increase in NODDI ICVF, with no concomitant change in ISOVF, is unclear, but could partly be attributed to axonal swelling (Moll et al., 2011), axonal bundling or changes in cytoskeleton composition following demyelination (Brady et al., 1999), or axonal repair. Axonal regeneration *per se* appears unlikely given the limited ability of the CNS to repair following axonal injury (Huebner and Strittmatter, 2009), and the lack of positive association with change in whole brain volume, an established marker of neurodegeneration. A decrease in glial cell infiltration following resolution of acute inflammation could additionally explain an increase in ICVF relative to a resulting decrease in hindered diffusion. Previous evidence also suggests that the parallel diffusivity in supratentorial brain may increase over time in RRMS (Harrison et al., 2011), which would artificially increase ICVF (Guerrero et al., 2019). The possibility that the assumption of a fixed intrinsic parallel diffusivity does not hold in NAWM can also not be excluded here.

The observed absence of longitudinal change in NODDI ISOVF was expected as significant increases in water content would likely be visible as hyperintense signal on conventional T2 FLAIR.

#### 4.2.3 g-ratio

The longitudinal increase in g-ratio within NAWM observed is also consistent with subtle demyelination in early RRMS which is not otherwise visible on conventional MRI. The MRI g-ratio has been studied in cross-sectional studies of MS (Kamagata et al., 2019; Maekawa et al., 2022; York et al., 2021; Yu et al., 2019), healthy cohorts (Mohammadi et al., 2015), other diseases (e.g. Moyamoya disease (Hara et al., 2020), Huntington’s disease (Johnson et al., 2021b)), and childhood development (Geeraert et al., 2019); however longitudinal analysis of g-ratio in the adult brain has been notably absent. The data presented illustrate that g-ratio may be suited to assessing myelin integrity changes over time. Conversely, an increase in g-ratio could also result from the observed increase in NODDI ICVF. Nevertheless, the strong relationship in NAWM between longitudinal change in g-ratio and MTsat but little association with NODDI measures suggests that myelin loss is driving the longitudinal increase in NAWM g-ratio.

The majority of patient data points for g-ratio in NAWM fell within limits of agreement established from healthy control test-retest measures. While the significant group-wise longitudinal change in g-ratio indicates a biological change in NAWM over one year post-diagnosis in RRMS, such change may not exceed measurement error for an individual patient.

### 4.3 Longitudinal microstructural changes in WML

#### 4.3.1 MTI

In WML, MTsat increased over one year, suggestive of myelin repair. As far as these authors are aware, this is the first study to examine longitudinal evolution of MTsat in MS lesions. The findings presented are, however, in line with previous post-mortem evidence showing that the percentage of remyelinated tissue in WML may be as high as 85% in RRMS, although highly heterogeneous across patients (Patrikios et al., 2006). Furthermore, WML MTsat at follow-up remained lower than NAWM values, in keeping with reports that myelin sheaths of remyelinated axons within T2 WML are abnormally thin (Barkhof et al., 2003) and remyelination is patchy (Patrikios et al., 2006). Although a decrease in water content could also account for a lesion-specific increase in MTsat, the increase in NODDI ISOVF within lesions, and the previously reported lack of change in total water content in existing WML (Vavasour et al., 2021), suggests this is not the case. Moreover, the effect size was large with several patients exceeding limits of agreement established in control white matter. Some recovery of myelin following acute inflammation therefore appears to be a plausible explanation.

Early treatment with DMTs, which target inflammation and reduce likelihood of progression (Brown et al., 2019), could also contribute to the increase in WML MTsat. Our cohort was treatment-naïve at baseline but nearly two thirds of patients had commenced DMTs by one year follow-up. Initiation of DMTs was not a significant covariate in WML models, however, suggesting that spontaneous remyelination may be a more significant effect. Reclassification of NAWM tissue as WML at follow-up is a potential additional explanation for an increase in MTsat over time, as new lesions may not be as extensively damaged. Our comparison of patients with and without new lesions at follow-up, however, showed a similar effect in both groups.

Unlike MTsat, MTR in WML did not change significantly post-diagnosis, after accounting for confounding variables including lesion load and age. The relative stability of MTR within WML has been noted previously (York et al., 2022), although MTR may fluctuate with time and lesion type (e.g. contrast-enhancing versus non-enhancing lesions) (Brown et al., 2014; York et al., 2022). Here, we did not investigate lesion sub-types; nonetheless, the discrepancy between results for MTsat and MTR suggests that T1 may be a contributing factor. The effect of T1 is likely to be greater in WML compared to NAWM, where marked myelin loss is associated with varying degrees of T1 prolongation, including visible black hole lesions in more extreme cases. Taken together, results suggest MTsat may be more sensitive to alterations in myelin integrity than MTR, including spontaneous remyelination.

#### 4.3.2 NODDI

The increase in NODDI ICVF within cerebral WML was greater than in NAWM and exceeded limits of agreement from healthy controls, although WML ICVF remained lower than in NAWM. Pathologically swollen axons are seen in secondary progressive MS (SPMS) lesions (Moll et al., 2011) and similar pathology occurring in early RRMS could explain our finding. A post-relapse reduction in glial cell presence, poor adaptability of the NODDI model to pathological tissue, residual sensitivity to remyelination, or alternatively axonal regeneration may be other explanations. The longitudinal increase in NODDI ISOVF within WML, however, does suggest ongoing progressive destruction of neuroaxonal architecture. This explanation would fit with the reduced axonal count typically seen in SPMS WML (Moll et al., 2011), but would seem to contradict myelin repair indicated by MTsat results. Heterogeneous tissue repair and destruction across/within lesions could perhaps consolidate these conflicting theories.

#### 4.3.3 g-ratio

The lack of change in g-ratio within WML may be due to competing effects; increase in MTsat, suggestive of remyelination, combined with significant increases in NODDI ICVF and NODDI ISOVF may mitigate each other, as discussed below. Intra-patient heterogeneity across lesions may also explain the negative result, and g-ratio changes on a lesion-by-lesion basis cannot be excluded.

### 4.4 g-ratio dependence on MTI and NODDI parameters

Simulation of the impact of concurrent changes in microstructural metrics on the g-ratio was performed *post hoc* to understand better the results obtained, and sheds light on the complexity of interpreting longitudinal change in MRI markers. As expected with demyelination in RRMS, decreasing MTsat leads to an increase in MRI g-ratio. Only a weak correlation between longitudinal changes in MTsat and g-ratio in WML, is however seen, despite a strong correlation in NAWM. While MTsat may be sensitive to myelinated axonal integrity, there may be a “floor” effect in focal regions of low myelin density which may limit the usefulness of the MTsat signal within WML.

Moreover, significant changes in NODDI measures are problematic for interpretation of the g-ratio. In RRMS, axonal density, measured here with NODDI ICVF, is expected to decrease with neurodegenerative processes and free water (i.e. ISOVF) is expected to increase as tissue destruction becomes more pronounced.

Simulations suggest, however, that a large increase in ISOVF, or a large decrease in ICVF, without a concomitant increase in MTsat would lead to a decrease in g-ratio (see Appendix Fig. A.3). While the latter scenario would not be expected in MS pathology given the proximity in time of demyelination and axonal degeneration, increases in ISOVF may be more common, particularly in WML; thus rendering the g-ratio model flawed in severely damaged WML. Nonetheless, the g-ratio may remain relevant in NAWM, where large changes in ISOVF and ICVF are not expected.

Although the g-ratio is clearly attractive as a parameter with a specific histopathological correlate and provides a mechanistic link between white matter integrity and neuronal conductivity in RRMS, these data suggest that g-ratio may not provide significant additional information to MTsat.

### 4.5 Limitations

There are a number of limitations in the present study. NODDI, MTsat and g-ratio analyses are heavily model-dependent and based on assumptions from healthy brain tissue, some of which may break down where there is marked loss of normal microstructural integrity. For example, the g-ratio model applied here does not account for the impact of varying neurite orientations on a sub-voxel level. NODDI ODI did not change significantly over one year, however, and was not associated with change in g-ratio (data not shown). Moreover, true evaluation of these techniques’ sensitivity to change in myelin and axonal integrity would require comparison with ‘ground truth’ brain tissue examination, which is not available in early RRMS, and a limitation common to the majority of imaging biomarker studies.

The cohort characteristics also impose limitations on analysis; over one year, a substantial number of patients will be recovering from an acute inflammatory episode, and progression in disability measures was minimal (Kearns et al., 2019), limiting the opportunity for correlating microstructural imaging measures with clinical progression. Future planned follow-up at five years will help to mitigate this limitation.

Although all participants were diagnosed within six months prior to baseline, the time duration between first symptom onset and diagnosis was variable, and not corrected for here due to an association with age (Spearman’s rho=0.48, p<0.001, data not shown). Nevertheless, longitudinal data from a sizable, comparatively homogeneous RRMS cohort who were recruited at a similar disease stage (Kearns et al., 2022) is a strength of this study.

Test-retest measures are important to establish sensitivity to detection of pathological change, however these were calculated from white matter in a small number of healthy control subjects and thus have wide confidence intervals, and may not be representative of WML.

Finally, loss of subject data to drop-out and technical inaccuracies may introduce bias to the analyses.

### 4.6 Conclusion

Measures specific to microstructural integrity show change in early RRMS where there is no detectable atrophy. MTsat is a promising *in vivo* biomarker of myelin integrity, which appears more sensitive than MTR to demyelination and spontaneous remyelination in early RRMS. G-ratio, despite its sensitivity to changes in NAWM and specific histopathological correlate with myelin thickness, is difficult to interpret biologically due to a complex dependence on NODDI parameters. Independent consideration of myelin-sensitive and axonal neuroimaging markers may ultimately be more informative for longitudinal tracking of neuropathology in RRMS than combining such measures. For clinical application, further research is required to improve technique reproducibility, broaden the applicability of dMRI and MTI models, and validate these against heterogeneous tissue pathology in MS.

## Data Availability

Data requests from a third party will be handled on a case by case basis by the steering committee and data custodians.

## Abbreviations

CNS: Central nervous system
dMRI: Diffusion-weighted magnetic resonance imaging
DMTs: Disease-modifying therapies
EDSS: Expanded Disability Status Scale
FLAIR: Fluid-Attenuated Inversion Recovery
FLASH: Fast Low-Angle Shot
ICVF: (NODDI-derived) intraneurite signal fraction
ihMTR: Inhomogeneous magnetisation transfer ratio
ISOVF: (NODDI-derived) isotropic signal fraction
MRI: Magnetic resonance imaging
MS: Multiple sclerosis
MTI: Magnetisation transfer imaging
MTR: Magnetisation transfer ratio
MTsat: Magnetisation transfer saturation
NAWM: ‘Normal-appearing’ white matter
NODDI: Neurite Orientation Dispersion and Density Imaging
ODI: (NODDI-derived) orientation dispersion index
RRMS: Relapsing-remitting multiple sclerosis
SNR: Signal-to-noise
SPMS: Secondary progressive multiple sclerosis
WML: White matter lesions

## Acknowledgements

With special thanks to all participants in FutureMS, the radiographers at the RIE Edinburgh, and Dr. Daisy Mollison. We would also like to thank other non-author contributors of the FutureMS consortium as follows: Chris Batchelor, Emily Beswick, Fraser Brown, Tracy Brunton, Jessie Chang, Yingdi Chen, Shuna Colville, Peter Connick, Annette Cooper, Denise Cranley, Rachel Dakin, Baljean Dhillon, Liz Elliott, Peter Foley, Stella Glasmacher, Angus Grossart, Haane Haagenrud, Katarzyna Hafezi, Emily Harrison, Sara Hathorn, David Hunt, Aidan Hutchison, Charlotte Jardine, Kiran Jayprakash, Matt Justin, Patrick Kearns, Lucy Kessler, Michaela Kleynhans, Juan Larraz, Dawn Lyle, Niall MacDougall, Jen MacFarlane, Alan Maclean, Bev MacLennan, Nicola Macleod, Don Mahad, Sarah-Jane Martin, Mary Monaghan, Lee Murphy, Katy Murray, Judith Newton, Julian Ng Kee Kwong, David Perry, Suzanne Quigley, Scott Semple, Adam Scotson, Amy Stenson, Maria Valdez Hernandez, Christine Weaver, Belinda Weller, Anna Williams, Stewart Wiseman, Charis Wong, Michael Wong and Rosie Woodward.

## Funding Sources

ENY was supported by a Chief Scientist Office SPRINT MND/MS Studentship and funding from the Anne Rowling Regenerative Neurology Clinic, Edinburgh. MJT is funded by the NHS Lothian Research and Development Office. RM and AK are funded by the UK MS Society Edinburgh Centre for MS Research grant (grant reference 133). FutureMS, hosted by Precision Medicine Scotland Innovation Centre (PMS-IC), was funded by a grant from the Scottish Funding Council to PMS-IC and Biogen Idec Ltd Insurance. Additional funding for the University of Edinburgh 3T MRI Research scanner in Royal Infirmary Edinburgh is funded by Wellcome (104916), Dunhill Trust (R380R/1114), Edinburgh and Lothians Health Foundation (2012/17), Muir Maxwell Research Fund, Edinburgh Imaging, and University of Edinburgh. For the purpose of open access, the author has applied a Creative Commons Attribution (CC BY) licence to any Author Accepted Manuscript version arising from this submission.

## Competing interests

The authors declare no conflicts of interests.

## Notes

### Competing Interest Statement

The authors have declared no competing interest.

### Clinical Protocols

https://wellcomeopenresearch.org/articles/7-94/v1

### Author Declarations

The South East Scotland Research Ethics Committee 02 of the National Health Service (NHS, United Kingdom) gave ethical approval for this work under reference 15/SS/023.

### Summary of Updates

Abstract and manuscript substantially shortened and restructured to focus on longitudinal aspects of study with clearer narrative. Supplemental files updated.

