## Appendix for "Longitudinal microstructural MRI markers of demyelination and neurodegeneration in early relapsing-remitting multiple sclerosis: magnetisation transfer, water diffusion and g-ratio"

Appendix A. Supplementary Material

| **Supplementary Tables** |  | Page |
| --- | --- | --- |
| Table A.1 | Linear mixed model results: g-ratio in NAWM | 2 |
| Table A.2 | Linear mixed model results: NODDI ICVF in NAWM | 3 |
| Table A.3 | Linear mixed model results: MTsat in NAWM | 4 |
| Table A.4 | Linear mixed model results: MTsat in WML | 5 |
| Table A.5 | Linear mixed model results: WML NODDI ICVF | 6 |
| Table A.6 | Linear mixed model results: NODDI ISOVF in WML | 7 |
| Table A.7 | Linear mixed model results: MTR in WML | 8 |
| Table A.8 | Linear mixed model results: whole brain volume | 9 |
| **Supplementary Figures** |  |  |
| Figure A.1 | Bland-Altman plots for MTI and NODDI metrics | 10 |
| Figure A.2 | Bland-Altman plots for MVF, AVF and g-ratio | 11 |
| Figure A.3 | Simulations showing the relationship between MVF, AVF and g-ratio | 12 |
| Figure A.4 | Boxplots showing longitudinal change in microstructural measures with healthy control limits of agreement | 13 |
| Figure A.5 | Scatterplots showing correlations between change over one year in ‘normal-appearing’ white matter for different metrics | 14 |
| Figure A.6 | Scatterplots showing correlations between change over one year in white matter lesions for different metrics | 15 |
| Figure A.7 | Scatterplots showing brain atrophy versus change in microstructural metrics | 16 |

Table A.1: Linear mixed model fit by maximum likelihood with mean g-ratio in normal-appearing white matter (NAWM) as the dependent variable (n=60). *significant after correction for FDR multiple comparisons

| **Random Effects** | **Estimate** |  |  |
| --- | --- | --- | --- |
| Subject (Intercept) | 0.0002 |  |  |
| Residual | 0.00007 |  |  |
| **Fixed Effects** | **Estimate** | **t-value (df)** | **p-value (uncorrected)** |
| (Intercept) | 0.561 | 71.33 (65.67) | <0.001 |
| Time | 0.005 | 3.08 (75.91) | 0.003* |
| Lesion load | 0.002 | 0.59 (71.04) | 0.555 |
| Age | 0.000 | 1.55 (63.06) | 0.127 |
| DMTs | -0.006 | -2.03 (74.04) | 0.046 |
| Sex | -0.003 | -0.57 (59.88) | 0.571 |
| **Conditional R^2^** | **Marginal R^2^** |  |  |
| 0.764 | 0.098 |  |  |

Table A.2: Linear mixed model fit by maximum likelihood with mean NODDI ICVF in cerebral normal-appearing white matter (NAWM) as the dependent variable (n=60).

| **Random Effects** | **Variance** |  |  |
| --- | --- | --- | --- |
| Subject (Intercept) | 0.0008 |  |  |
| Residual | 0.00002 |  |  |
| **Fixed Effects** | **Estimate** | **t-value (df)** | **p-value (uncorrected)** |
| (Intercept) | 0.5809 | 40.81 (58.07) | <0.001 |
| Time | 0.0035 | 3.51 (90.59) | 0.001* |
| Lesion load | -0.0128 | -3.51 (119.91) | 0.001 |
| Age | 0.0001 | 0.51 (60.77) | 0.613 |
| DMTs | -0.0003 | -0.19 (59.13) | 0.853 |
| Sex | 0.0073 | 0.83 (57.84) | 0.411 |
| **Conditional R^2^** | **Marginal R^2^** |  |  |
| 0.976 | 0.106 |  |  |

Table A.3: Linear mixed model for NAWM MTsat (n=62). *significant after FDR correction for multiple comparisons.

| **Random Effects** | **Variance** |  |  |
| --- | --- | --- | --- |
| Subject (Intercept) | 0.017 |  |  |
| Residual | 0.006 |  |  |
| **Fixed Effects** | **Estimate** | **t-value (df)** | **p-value (uncorrected)** |
| (Intercept) | 4.077 | 38.21 (87.97) | <0.001 |
| Time | -0.040 | -2.60 (79.26) | 0.011* |
| Lesion load | -0.228 | -2.70 (95.89) | 0.008 |
| Age | -0.007 | -2.61 (90.45) | 0.011 |
| DMTs | 0.067 | 2.50 (79.29) | 0.015 |
| Sex | 0.048 | 1.11 (62.13) | 0.270 |
| Lesion load * Age | 0.004 | 2.11 (102.79) | 0.037 |
| **Conditional R^2^** | **Marginal R^2^** |  |  |
| 0.795 | 0.211 |  |  |

Table A.4: Linear mixed model for WML MTsat (n=62). *significant after FDR correction for multiple comparisons.

| **Random Effects** | **Variance** | | |  |  |
| --- | --- | --- | --- | --- | --- |
| Subject (Intercept) | 0.0695 | | |  |  |
| Residual | 0.0052 | | |  |  |
| **Fixed Effects** | **Estimate** | | | **t-value (df)** | **p-value (uncorrected)** |
| (Intercept) | 2.178 | | | 12.46 (99.37) | < 0.001 |
| Time | 0.059 | | | 3.65 (82.58) | < 0.001* |
| Lesion load | 0.266 | | | 2.11 (123.89) | 0.037 |
| Age | 0.006 | | | 1.32 (107.61) | 0.189 |
| DMTs | -003 | | | -0.10 (68.41) | 0.923 |
| Sex | -0.083 | | | -1.00 (61.77) | 0.321 |
| Lesion load * Age | -0.006 | | | -2.19 (120.29) | 0.031 |
| **Conditional R^2^** | | **Marginal R^2^** |  | | |
| 0.935 | | 0.064 |  | | |

Table A.5: Linear mixed model fit by maximum likelihood with mean NODDI ICVF in cerebral white matter lesions (WMLs) as the dependent variable (n=60). * significant after FDR correction for multiple comparisons

| **Random Effects** | **Variance** |  |  |
| --- | --- | --- | --- |
| Subject (Intercept) | 0.0013 |  |  |
| Residual | 0.0002 |  |  |
| **Fixed Effects** | **Estimate** | **t-value (df)** | **p-value (uncorrected)** |
| (Intercept) | 0.404 | 21.04 (59.44) | < 0.001 |
| Time | 0.017 | 6.95 (80.64) | < 0.001* |
| Lesion load | -0.003 | -0.40 (83.68) | 0.688 |
| Age | -0.0003 | -0.68 (61.90) | 0.498 |
| DMTs | -0.006 | -1.12 (65.30) | 0.267 |
| Sex | 0.008 | 0.70 (59.31) | 0.488 |
| **Conditional R^2^** | **Marginal R^2^** |  |  |
| 0.892 | 0.092 |  |  |

Table A.6: Linear mixed model fit by maximum likelihood with mean NODDI ISO in cerebral white matter lesions (WMLs) as the dependent variable (n=60). *significant after FDR-correction for multiple comparisons

| **Random Effects** | **Variance** |  |  |
| --- | --- | --- | --- |
| Subject (Intercept) | 0.0003 |  |  |
| Residual | 0.0001 |  |  |
| **Fixed Effects** | **Estimate** | **t-value (df)** | **p-value (uncorrected)** |
| (Intercept) | 0.064 | 6.39 (60.05) | <0.001 |
| Time | 0.011 | 5.56 (74.85) | <0.001* |
| Lesion load | -0.015 | -4.43 (69.70) | <0.001 |
| Age | 0.001 | 5.46 (63.29) | <0.001 |
| DMTs | -0.009 | -2.16 (76.25) | 0.034 |
| Sex | -0.006 | -0.90 (59.88) | 0.373 |
| **Conditional R^2^** | **Marginal R^2^** |  |  |
| 0.815 | 0.379 |  |  |

Table A.7: Linear mixed model fit by maximum likelihood with mean MTR in white matter lesions (WMLs) as the dependent variable (n=62). *significant after FDR correction for multiple comparisons.

| **Random Effects** | **Variance** | | |  |  |
| --- | --- | --- | --- | --- | --- |
| Subject (Intercept) | 3.077 | | |  |  |
| Residual | 0.500 | | |  |  |
| **Fixed Effects** | **Estimate** | | | **t-value (df)** | **p-value (uncorrected)** |
| (Intercept) | 44.928 | | | 34.60 (95.04) | <0.001 |
| Time | 0.241 | | | 1.60 (79.48) | 0.113 |
| Lesion load | 2.809 | | | 2.82 (113.49) | 0.006 |
| Age | 0.069 | | | 2.03 (101.06) | 0.044 |
| DMTs | 0.112 | | | 0.44 (72.30) | 0.664 |
| Sex | -0.880 | | | -1.56 (61.66) | 0.124 |
| Lesion load * Age | -0.065 | | | -2.71 (120.78) | 0.008 |
| **Conditional R^2^** | | **Marginal R^2^** |  | | |
| 0.876 | | 0.110 |  | | |

Table A.8: Linear mixed model results for whole brain volume (as a percentage of intracranial volume) (n=62). *significant after FDR correction for multiple comparisons

| **Random Effects** | **Estimate** |  |  |
| --- | --- | --- | --- |
| Subject (Intercept) | 0.0008 |  |  |
| Residual | 0.00002 |  |  |
| **Fixed Effects** | **Estimate** | **t-value (df)** | **p-value (uncorrected)** |
| (Intercept) | 0.813 | 59.93 (60.30) | <0.001 |
| Time | -0.001 | -0.91 (86.28) | 0.366 |
| Lesion load | -0.001 | -0.30 (123.79) | 0.763 |
| Age | -0.001 | -3.52 (61.38) | 0.001 |
| DMTs | -0.003 | -1.94 (60.49) | 0.057 |
| Sex | -0.013 | -1.56 (59.14) | 0.124 |
| **Conditional R^2^** | **Marginal R^2^** |  |  |
| 0.978 | 0.185 |  |  |

Figure A.1: Bland-Altman plots show test-retest agreement in white matter for healthy controls for (A) magnetisation transfer saturation and (B) magnetisation transfer ratio (%, n=11); and, (C) NODDI intracellular volume fraction and (D) NODDI isotropic volume fraction (n=9). NODDI measures do not include cerebellum due to technical inaccuracies.

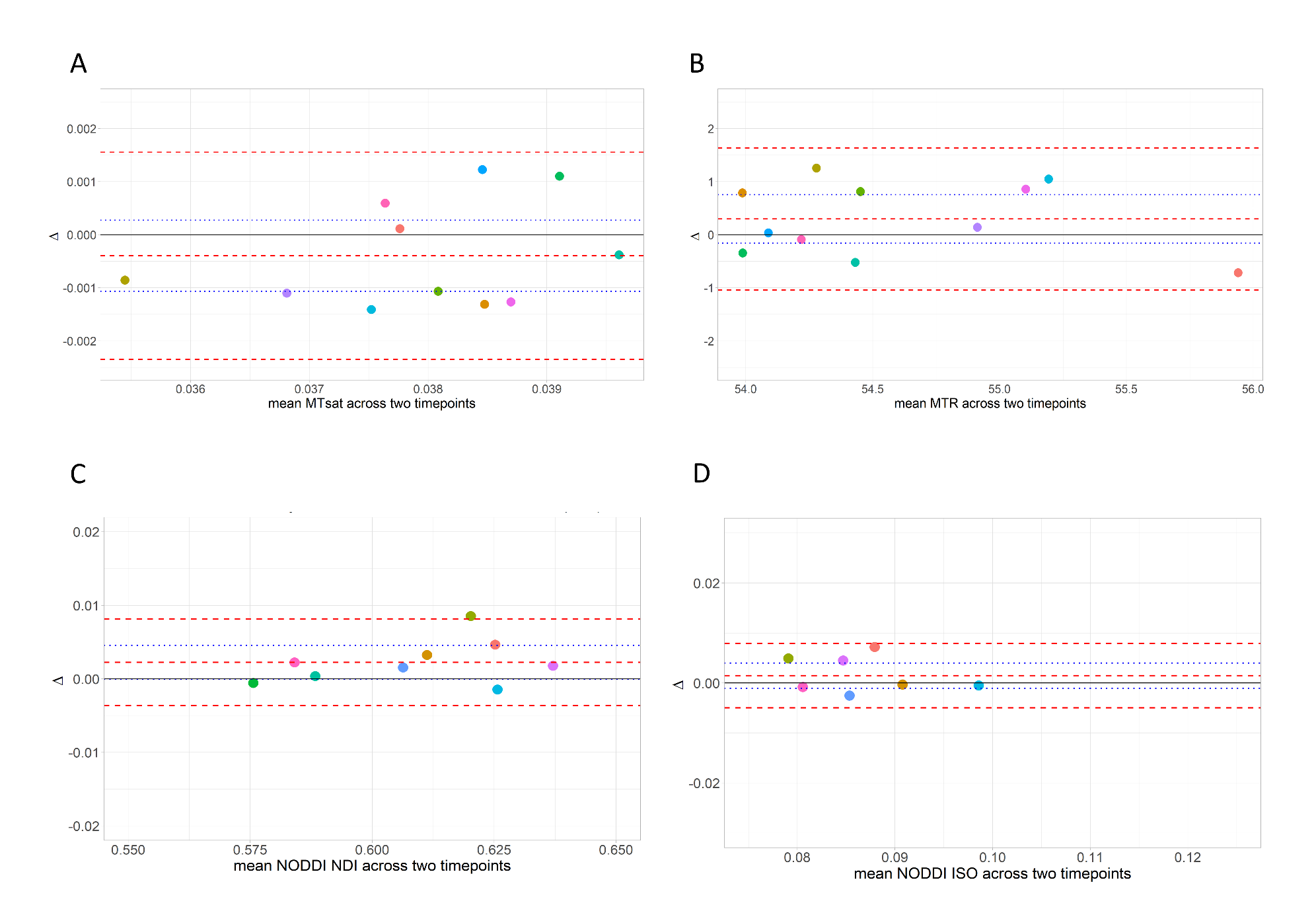

Figure A.2: Bland-Altman plots show test-retest agreement in healthy control white matter (n=9) for (A) myelin volume fraction, (B) axonal volume fraction, and (C) g-ratio.

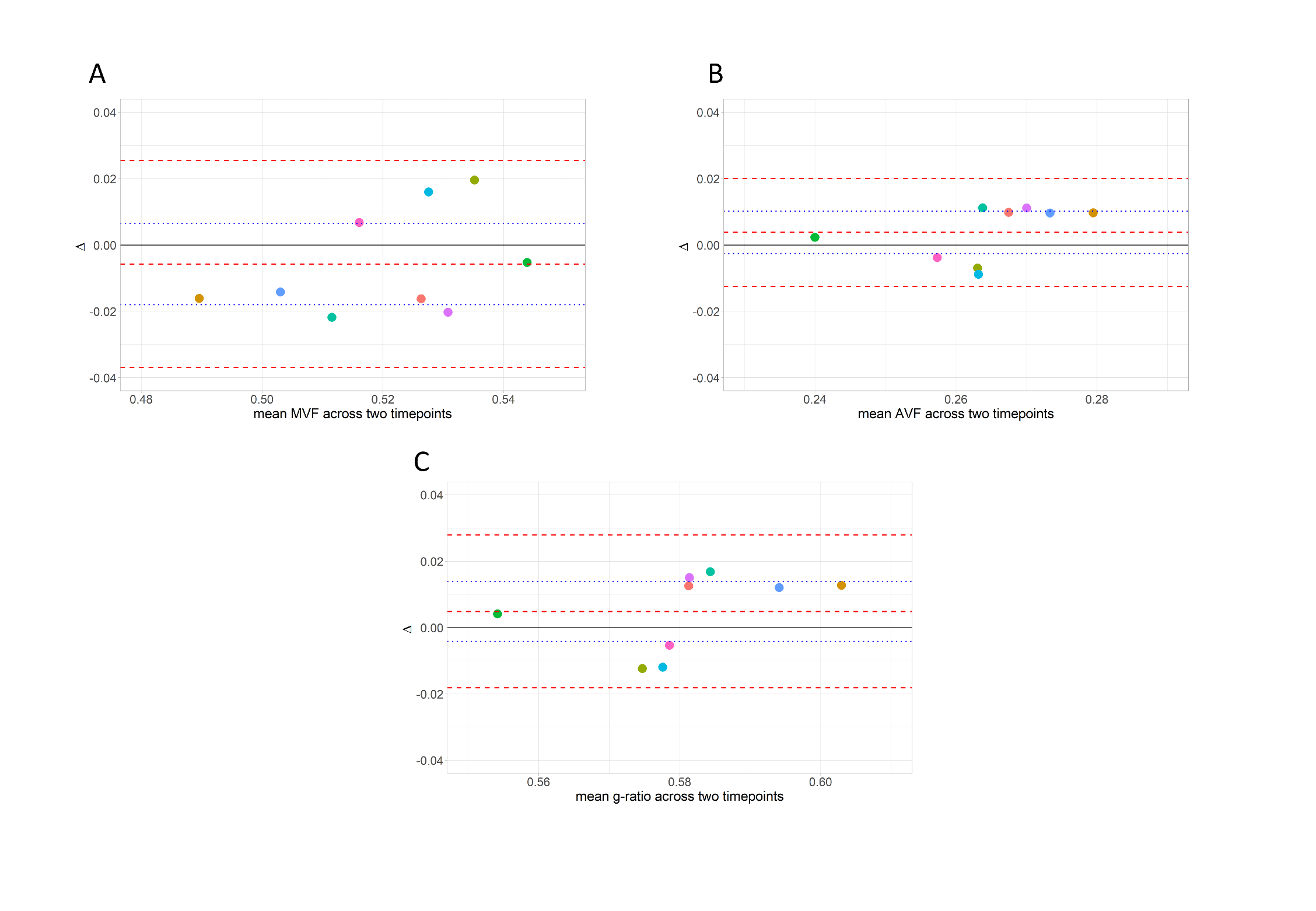

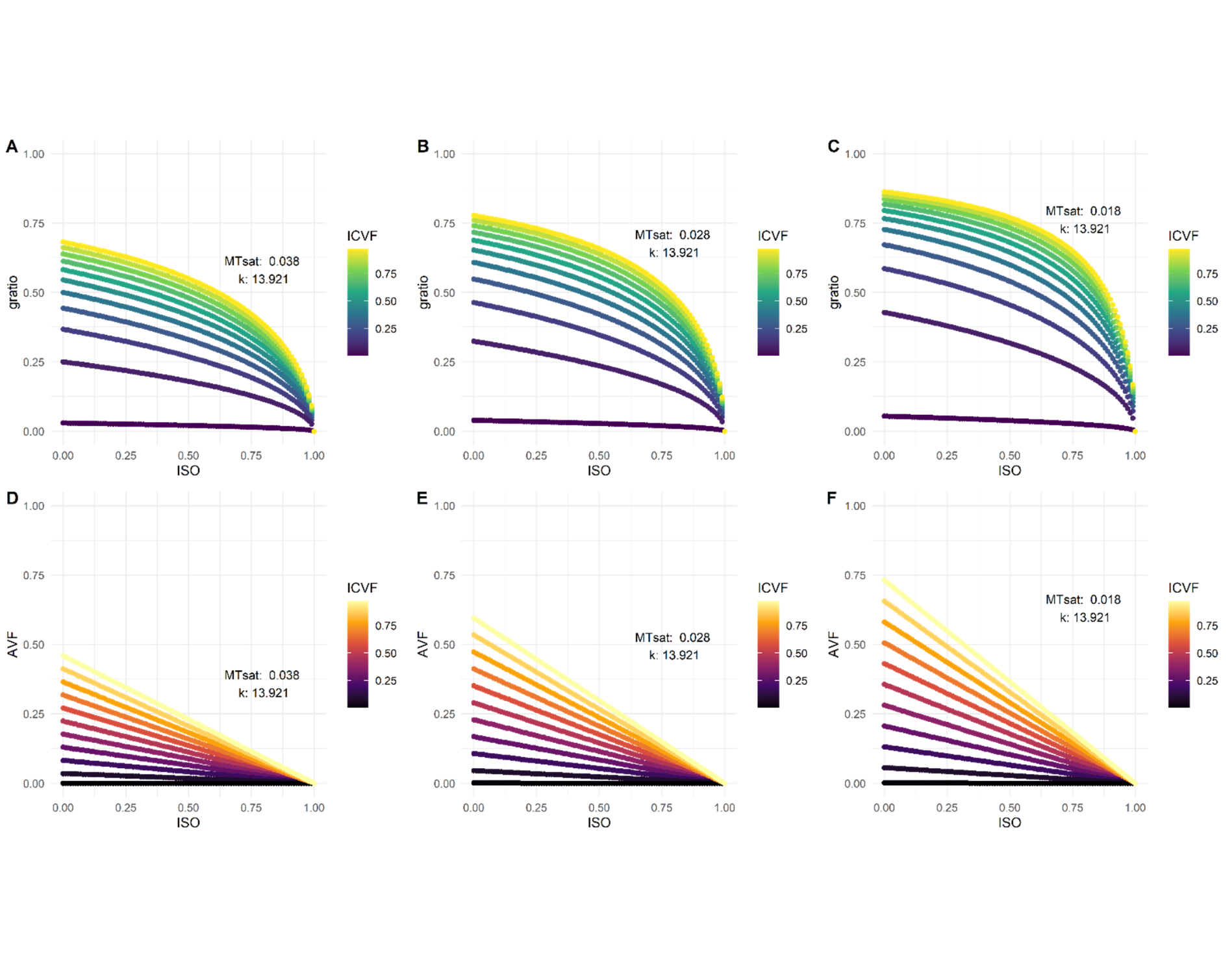

Figure A.3: Simulations show how g-ratio and the axonal volume fraction (AVF) would be expected to change with changes in magnetisation transfer saturation (MTsat) and NODDI isotropic and intraneurite volume fractions (ISO and ICVF, respectively). k: calibration constant.

Figure A.4: Boxplots show longitudinal change in normal-appearing white matter (NAWM) and white matter lesions (WMLs) for (A) MTsat, (B) MTR, (C) NODDI ICVF, (D) NODDI ISOVF and (E) g-ratio. Limits of agreement from healthy control white matter are superimposed (red dashed lines) with 95% confidence intervals (dotted lines).

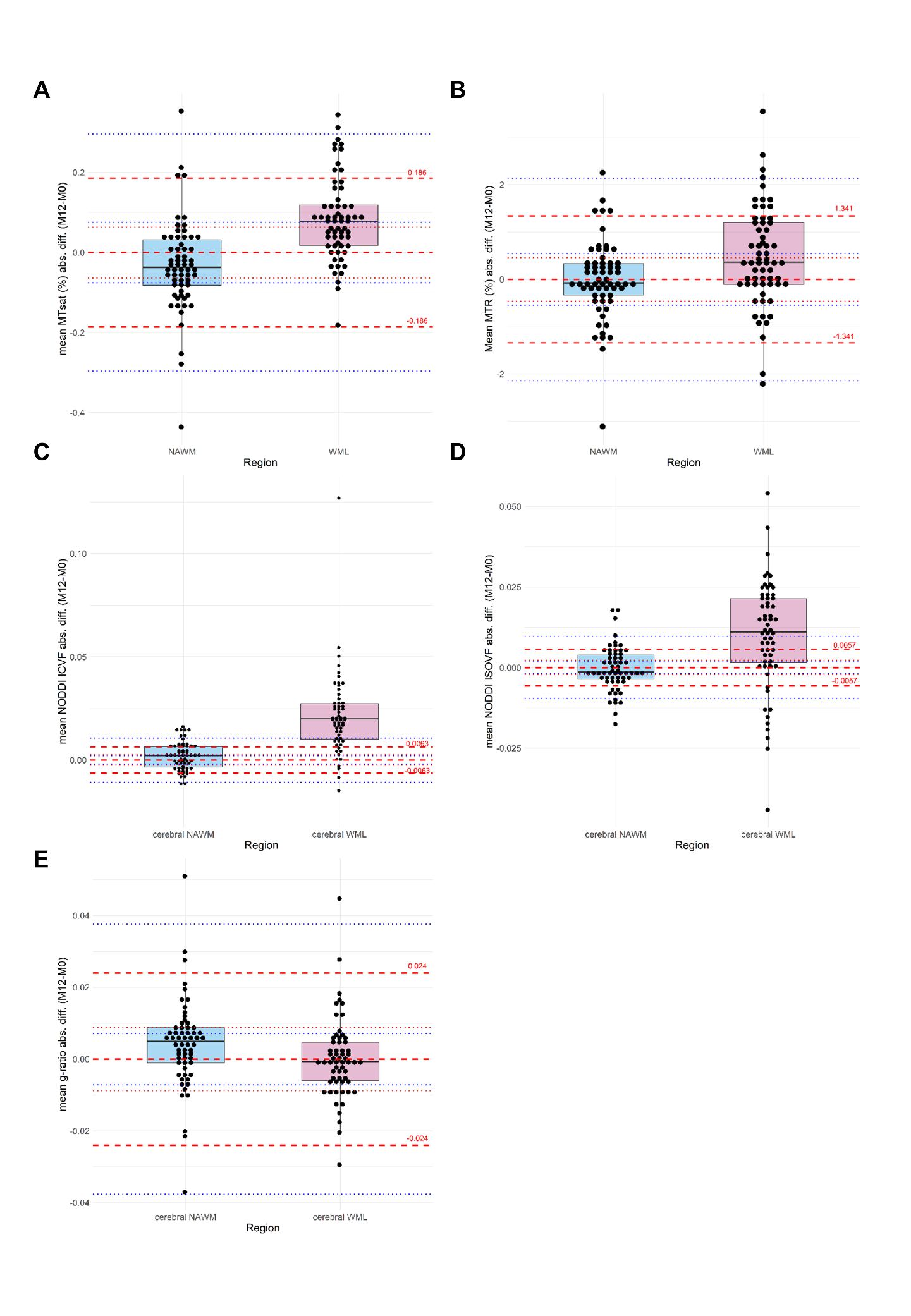

Figure A.5: Pearson's correlations between change over one year in ‘normal-appearing’ white matter g-ratio and (a) MTsat (%), (b) NODDI ICVF, (c) NODDI ISOVF; and between MTsat and (d) NODDI ICVF, and (e) NODDI ISOVF; and (f) NODDI ICVF and NODDI ISOVF.

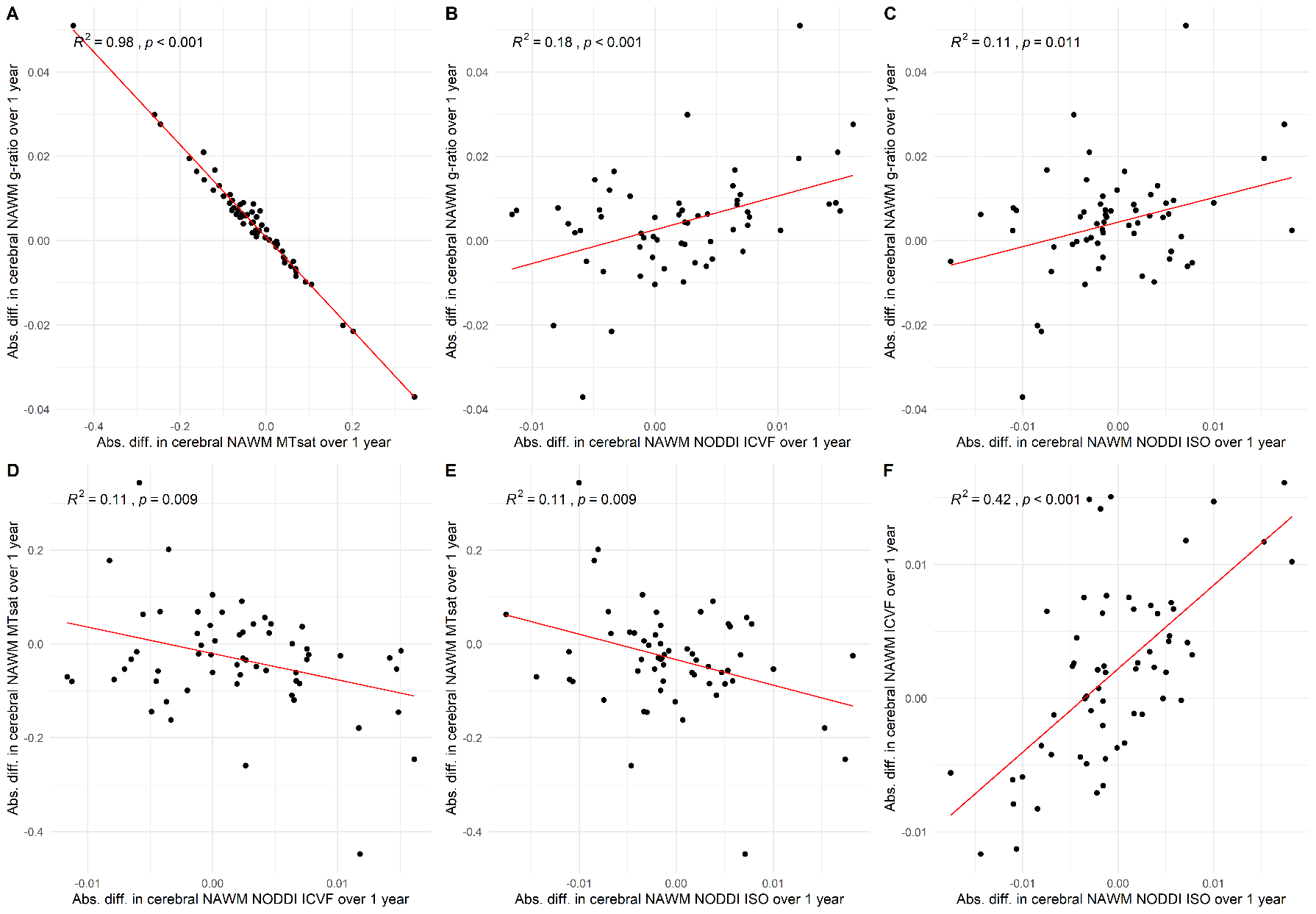

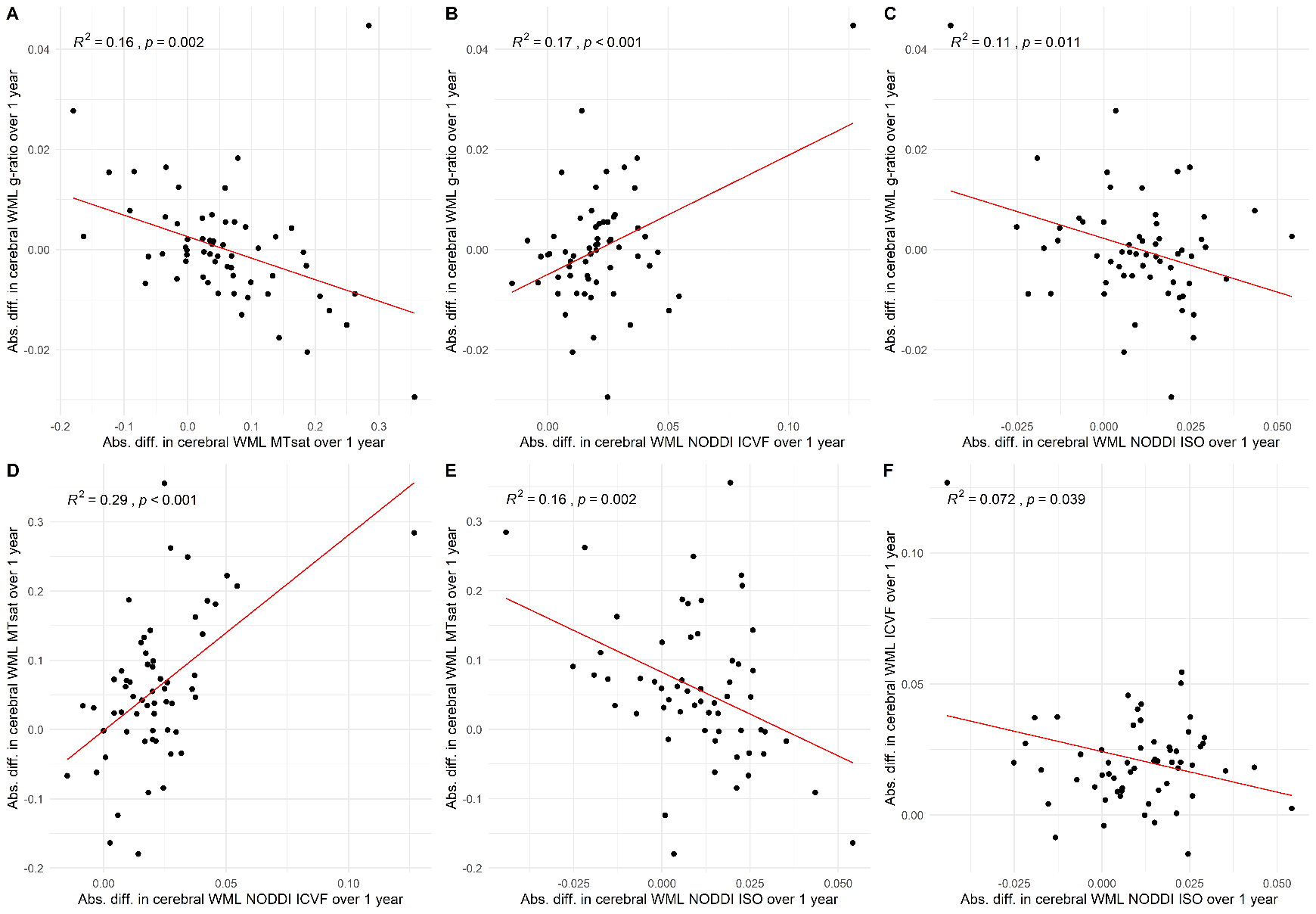

Figure A.6: Pearson's correlations between change over one year in WMLs between g-ratio and (a) MTsat (%), (b) NODDI ICVF, (c) NODDI ISO, and MTsat and (d) NODDI ICVF, and (e) NODDI ISO, and (f) NODDI ICVF and NODDI ISO.

Figure A.7: Scatterplots show change in whole brain volume over one year versus change in NAWM (A) MTsat (%), (B) NODDI ICVF, and (C) g-ratio, and versus change in WML (D) MTsat, (E) NODDI ICVF and (F) NODDI ISOVF.

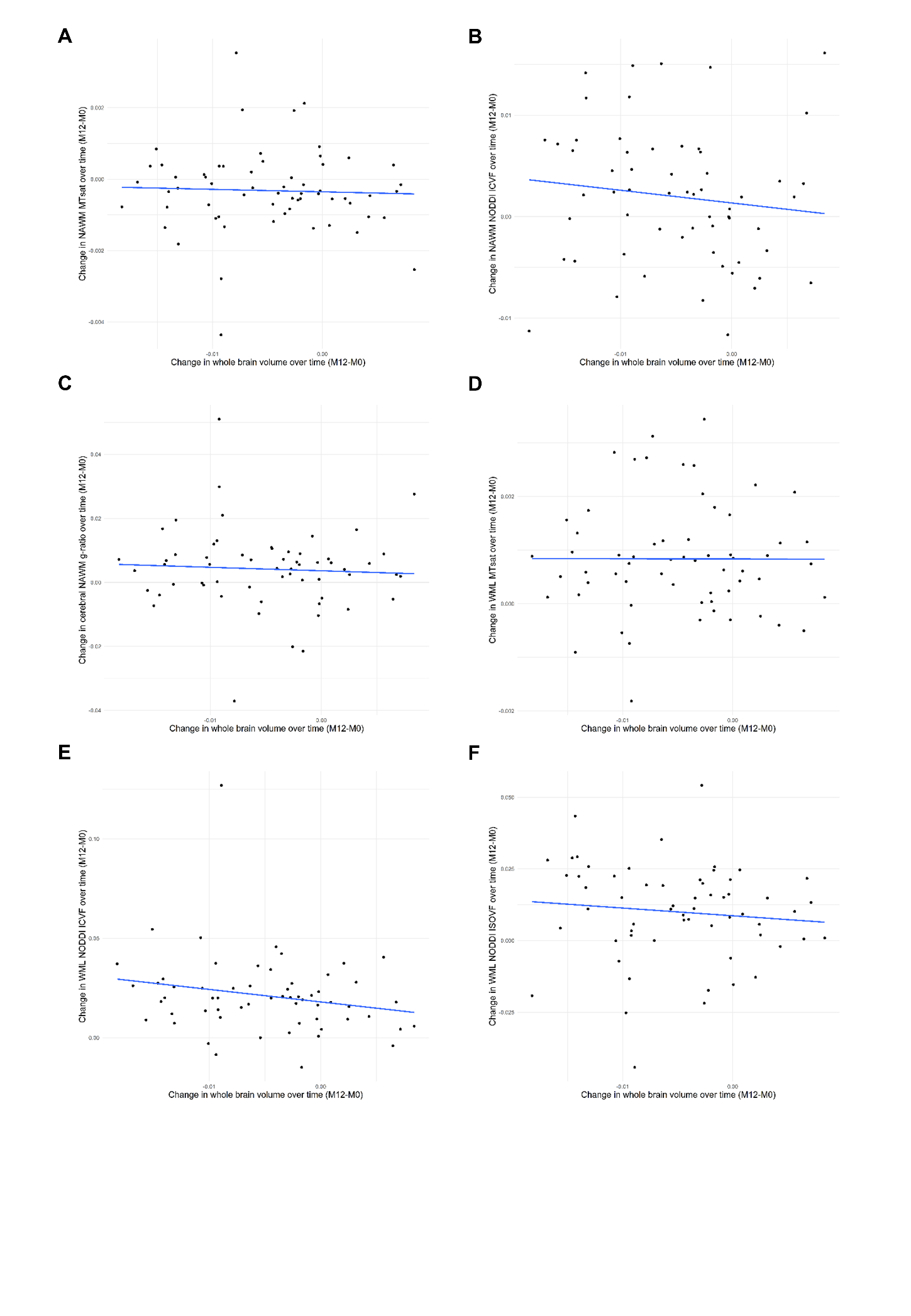
